# Classification of the Attempted Arm and Hand Movements of Patients with Spinal Cord Injury Using Deep Learning Approach

**DOI:** 10.1101/2023.07.06.23292320

**Authors:** Sahar Taghi Zadeh Makouei, Caglar Uyulan

## Abstract

The primary objective of this research is to improve the average classification performance for specific movements in patients with cervical spinal cord injury (SCI). The study utilizes a low-frequency multi-class electroencephalography (EEG) dataset obtained from the Institute of Neural Engineering at Graz University of Technology. The research combines convolutional neural network (CNN) and long-short-term memory (LSTM) architectures to uncover strong neural correlations between temporal and spatial aspects of the EEG signals associated with attempted arm and hand movements.

To achieve this, three different methods are used to select relevant features, and the proposed model’s robustness against variations in the data is validated using 10-fold cross-validation (CV). Furthermore, the study explores the potential for subject-specific adaptation in an online paradigm, extending the proof-of-concept for classifying movement attempts.

In summary, this research aims to make valuable contributions to the field of neuro-technology by developing EEG-controlled assistive devices using a generalized brain-computer interface (BCI) and deep learning (DL) framework. The focus is on capturing high-level spatiotemporal features and latent dependencies to enhance the performance and usability of EEG-based assistive technologies.

## 1. Introduction

Individuals experiencing spinal cord injury (SCI) commonly exhibit primary symptoms associated with the loss of motor and sensory functions. SCI is characterized by the disruption of specific neuronal pathways that connect the brain to the limbs, resulting in damage to the brain regions responsible for controlling these limbs [Lopez-Larraz et al., 2012]. This condition has significant physical and psychological consequences, necessitating substantial lifestyle adjustments. SCI can have devastating effects, including the loss of motor and sensory function, chronic pain syndromes, heightened susceptibility to depression, anxiety, and substance abuse, as well as increased hospitalizations and an overall decline in health. Trauma or illness-induced damage to the spinal cord leads to the demise of neural cells and debilitating sensory and motor function loss [Wei et al., 2009].

Individuals with mobility impairments caused by various conditions such as amyotrophic lateral sclerosis, brainstem trauma, brain or spinal cord injury, cerebral palsy, muscular dystrophies, and multiple sclerosis are unable to effectively interact with computers and machinery due to their compromised movement abilities. One approach to enable communication between these individuals and machines is through the identification and interpretation of brain activity patterns [Karakaya et al., 2017].

Brain-computer interfaces (BCIs) based on electroencephalograms (EEG) have garnered significant attention as a direct and non-invasive means of communication between external devices and the human brain. This method can be employed with almost any individual possessing a healthy brain and is safe, straightforward to implement, and non-invasive. The field of neuro-engineering has made promising advancements over the past two decades, demonstrating the potential use of BCIs in enhancing functional recovery and autonomy in individuals with motor impairments. BCIs bypass the impaired neuromotor system by converting brain activity into control signals for computers and machines. In recent years, BCIs have emerged as an alternative form of communication between the human brain and output devices. By capturing electrical signals from the scalp, the intentions of the user can be decoded in real time and translated into control commands for operating external devices, computer screens, and virtual objects. The application of BCIs involves identifying suitable physical and mental tasks, placing electrodes corresponding to these tasks, extracting relevant features from the recorded signals, developing high-performance classification algorithms, and transmitting this information to communication and control units via transducer algorithms. BCIs detect changes in brain activity when users move or intend to move, and translate these changes into control signals for neuroprostheses or robotic arms. BCI systems typically comprise signal acquisition and preprocessing, feature extraction, classification, and a control interface [Xu et al., 2022; Agarwal et al., 2020; Robinson et al., 2021; Tariq et al., 2018; Aydin et al., 2022]. Each BCI progresses through five primary stages: brain activity measurement, data preprocessing, feature extraction, classification, and the control interface, where meaningful information is extracted from the classified data for output devices.

As seen in *Fig.1*, in Human-Computer Interface systems, data received from humans are processed and transmitted to the computer environment. This environment may be an electronic device or a mechanical arm. In the signal processing portion, an accurate determination may be made through preprocessing, feature extraction, and classification.

**Figure 1:**
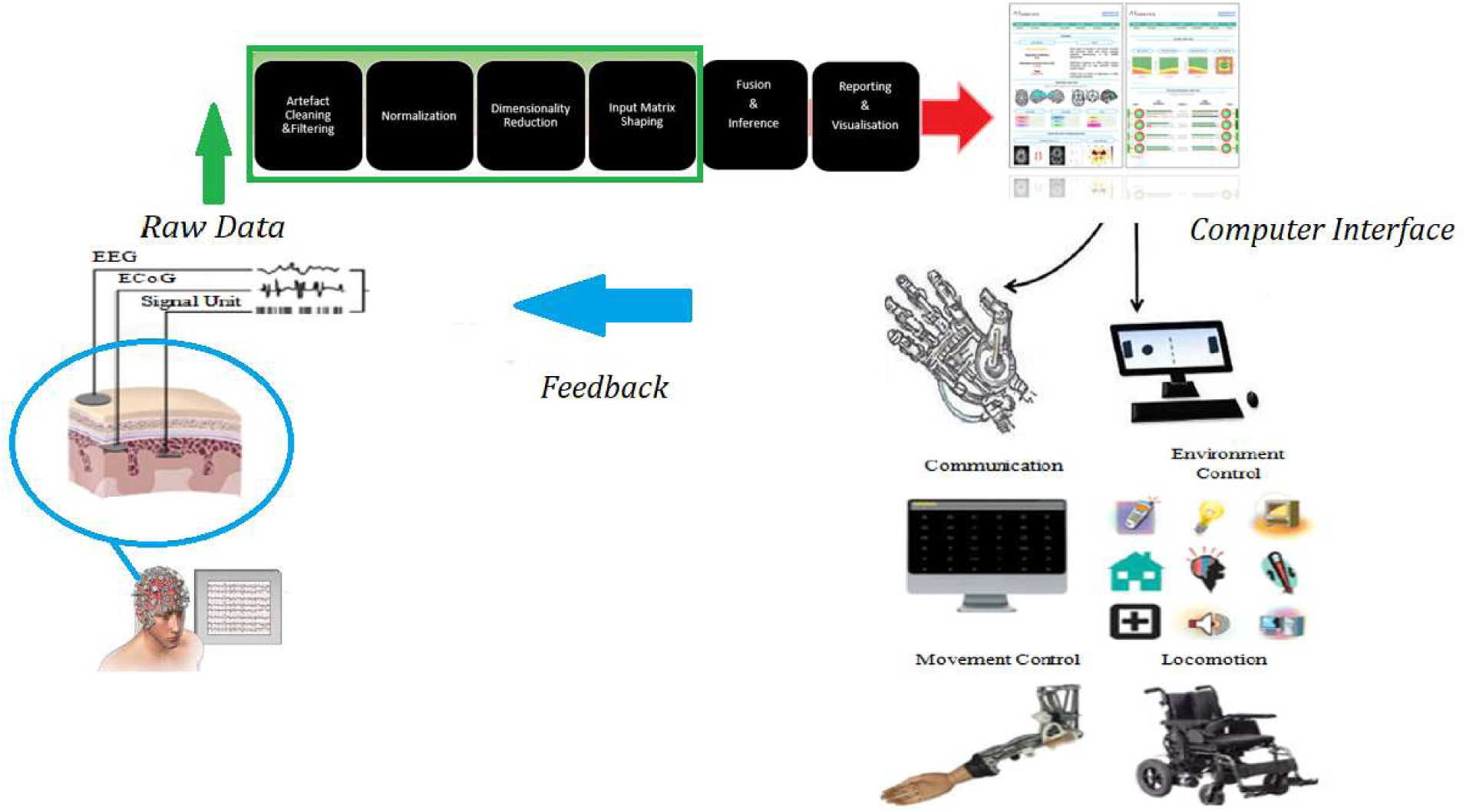
Human-computer interface system signal processing scheme [Mudgal et al., 2020; Kareem Abdullah& Chao Zhu, 2014].

EEG is utilized in BCI research due to its non-invasive nature, practicality, real-time capabilities, and wearable options. EEG signals, which are among the most favored methods for measuring electrical signals generated as a result of brain activity, are obtained through the use of electrodes placed on the user’s scalp. The high time resolution, painlessness, low cost in comparison to other methods, availability of wearable types, and harmlessness make it a preferred choice in BCI studies. Affordable devices are available for capturing brain signals, which serve as inputs for systems that decode the relationship between experimental arm and hand movements and variations in electroencephalographic (EEG) signals. These devices are known as EEG-based brain-computer interfaces (BCIs). The overarching goal of research in the field of EEG-based BCIs is to develop a method with higher classification rates and brain-computer interface data rates compared to existing methods [Aydemir & Kayikcioglu, 2014; Torres et al., 2020].

Electroencephalography (EEG) provides a powerful means of studying cortical plasticity. One of the main applications of EEG is monitoring brain activity associated with cognitive and movement-related processes. When individuals with complete spinal cord injuries (SCIs) are instructed to attempt movement with their paralyzed limbs, EEG data, or brain waves, represent the electrical signals generated by the excitatory and inhibitory potentials of neurons. The processing of EEG signals involves various aspects of noise rejection and feature vector extraction. Independent Component Analysis (ICA) is the most commonly used method for noise removal. Recently, a multivariate spectral technique called Directed Transfer Function (DTF) has been proposed to determine directional effects between different channels in multivariate datasets. Fast Fourier Transforms (FFT) is an algorithm used to transform data into the frequency domain, providing magnitude values (μV) for each frequency band. When comparing SCI and healthy control groups, magnitude difference datasets are generated for each frequency range. EEG signals offer excellent temporal resolution and directly measure neuronal activity. These signals cannot be manipulated or simulated to mimic hand movements, making them reliable sources of information. EEG-based BCI devices, such as non-invasive, low-cost, and wearable options like helmets and headbands, provide a cost-effective and convenient means of detecting EEG signals. Brain electrophysiological signals, including EEG and evoked potentials (EP), contain valuable information about the physiological states and functional activities of the brain. EEG activity is often characterized by five different frequency bands: delta (0.1 - 3 Hz), theta (4 - 7 Hz), alpha (8 - 13 Hz), beta (14 - 29 Hz), and gamma. Investigating alterations in the 8-13 Hz range of brain activity following an SCI is crucial, as studies suggest cortical remodeling that may impact activity within this frequency range [Stewart et al., 2014; Xu et al., 2022; Wei et al., 2009; Xu et al., 2022; Selvi et al., 2021].

Oscillatory activity, including delta, theta, alpha/mu, beta, and gamma rhythms, event-related potentials (ERPs) such as P300, visual evoked potentials (VEPs), and slow cortical potentials (SCPs), are important components for effective communication in EEG-based BCIs. Oscillating rhythms fluctuate based on the state of brain activity, with specific rhythms associated with particular situations. Mu and beta rhythms, referred to as sensorimotor rhythms (SMRs), exhibit event-related desynchronization (ERD) and event-related synchronization (ERS), respectively, which are directly linked to power decrease during movement execution or power increase during rest. ERPs, on the other hand, are phase-locked signals. Users can be categorized based on their physical and mental status; for example, locked-in patients with intact eye muscles can communicate via ERP signals, while individuals with motor-complete but sensory-deficient SCIs can utilize SMR-based motor imagery signals. EEG signals provide rich information such as emotional, motor, and visual states. However, EEG signals are complex and require feature extraction techniques to obtain the desired signal structure. As non-stationary signals containing a large amount of data, EEG signals consist of spikes and field potentials. Spikes represent individual action potentials of neurons and are detected using invasive microelectrodes, while field potentials can be measured by EEG and reflect the combined synaptic, axonal, and neuronal activity of groups of neurons. EEG signals are obtained by measuring the currents resulting from synaptic stimulation, which create a magnetic field that can be measured by an EEG device. Electrodes are attached to different parts of the brain to measure and record this magnetic field, and the impedance of the connection electrodes should be chosen accordingly due to the differing impedances at each brain stage. EEG signals are recorded after appropriately placing electrodes on the human skull, typically using multiple channels to capture signals more accurately. Downsampling and filtering are applied to address noise issues in the EEG signals, with Butterworth filtering commonly used. The combination of downsampling and filtering reduces training time for model algorithms, resulting in efficient and fast models. A comparison of EEG signals before and after downsampling and filtering is presented.

### Related Works

A novel approach called Proto-imEEG, based on a Prototypical Network, has been developed for automatically classifying imagined speech prompts and words using EEG data. This approach incorporates a one-dimensional convolutional layer and bidirectional recurrent networks, surpassing the performance of state-of-the-art methods with an average classification accuracy of 92-96% and an average inference time of 0.2 seconds. This study represents the first application of a Prototypical Network in this context [Hernandez-Galvan et al., 2022].

Three hybrid models combining convolutional neural networks (CNNs) with Long-Short Term Memory (LSTM) networks were proposed for classifying EEG signals in motor imagery-based BCIs. The models were evaluated using the BCI Competition IV dataset 2a, and the hybrid neural network with Inception-v3 achieved the highest mean accuracy of 92% and a mean Kappa value of 88%. These findings suggest that transfer learning, using a pre-trained CNN combined with LSTM, holds promise for motor imagery-based BCIs and has the potential to reduce computational time by selecting the most discriminative channels for different motor imagery tasks in distinct brain regions [Khademi et al., 2022].

Accurate classification of target movements in EEG and EMG signals is crucial for effective control of prosthetics. However, missing data in these signals can significantly decrease classification accuracy and impair prosthetic control. A framework combining tensor factorization and an attention-based CNN-LSTM deep learning method was proposed to recover missing data and classify target movements, respectively. The tensor factorization employed Canonical/Polyadic Weighted OPTimization (CP-WOPT), and the CNN-LSTM-Attn classifier demonstrated mean classification accuracies of 98%, 83%, and 90% on complete, partially complete, and tensor-recovered real-world EEG and EMG data, respectively, indicating its effectiveness in this application [Akmal, 2022].

A hybrid deep learning method, combining a one-dimensional CNN and a long short-term memory (LSTM) model, was employed for the classification of four motor imagery tasks using EEG and electrooculogram (EOG) data. The proposed method showed improved classification accuracy compared to state-of-the-art methods and exhibited robustness against data variations. The model’s performance was evaluated using various metrics, including accuracy, kappa value, receiver operator characteristic curve, and area under the curve [Uyulan, 2021].

A hybrid deep learning approach was presented for classifying four-class motor imagery EEG signals. The proposed algorithm utilized a filter bank common spatial pattern for feature extraction and a hybrid deep network consisting of a convolutional neural network and a long-term short-term memory network to learn spatial and temporal features simultaneously. The shared neural network, trained using data from all subjects, achieved an accuracy of 83% and a Cohen’s kappa value of 0.80 on a brain-computer interface competition dataset. Additionally, the shared neural network demonstrated satisfactory accuracy when evaluated on a subject-by-subject basis [Zhang et al., 2019].

A model for identifying EEG signals of children with attention deficit hyperactivity disorder (ADHD) was presented, consisting of temporal convolutional blocks, spatial convolutional blocks, and an LSTM layer. This model, designed to extract more temporal information from the EEG data, outperformed Support Vector Machine (SVM) and EEGNet in terms of correct classification rates. The model achieved correct classification rates of 92.29%, 92.76%, and 90.91% on the three backs, respectively, and 94.25% on the full dataset, demonstrating its feasibility and applicability to ADHD recognition [Zhang et al., 2022].

The development of a BCI system for controlling the movement of a hexapod robot using fist motor imagery signals was presented. The system utilized EEG signals sensed on the human sensorimotor cortex to classify motor imagery EEG signals and identify commands for controlling the robot’s locomotion. A hybrid architecture combining CNNs and LSTM networks achieved accuracies of 84.69% and 79.2% on two datasets, respectively, demonstrating the feasibility of using motor imagery EEG signals in BCI systems for controlling mobile robots and related applications [Mwata-Velu et al., 2021].

## 2. Material and Method

### 2.1.Experimental Design

We utilized EEG data from an online repository, collected from 10 participants with cervical spinal cord injury (SCI) within the age range of 20 to 69 years. The majority of participants were male, and all were originally right-handed. Two paradigms were employed: an offline paradigm involving 9 participants with a single session each, and an online paradigm with one participant and two sessions. In the offline paradigm, participants focused on a fixation cross and were instructed to avoid eye movements, blinking, and swallowing while executing or attempting hand movements corresponding to five different classes. This paradigm consisted of 9 runs, with each run comprising 40 trials, including eye movement and rest conditions. Two separate paradigms were used for training and evaluating a classifier for hand movements.

The training paradigm involved two types of trials: movement trials and rest trials. In a movement trial, participants were presented with a class cue indicating either hand open or palmar grasp, followed by a ready cue and then a go cue. Participants were instructed to perform the corresponding movement when the go cue appeared. In the rest trial, participants were instructed to refrain from any movement.

The test paradigm also included movement and rest trials. At the beginning of each trial, a class cue was presented, and participants attempted to perform multiple self-paced movements of the requested movement class for 60 seconds. Participants reported any movement attempts, and the experimenter marked the time point of each movement event. The online classifier remained active throughout the sessions and displayed the corresponding movement icon (hand open or palmar grasp) for 2 seconds whenever a movement attempt was detected.

The study recorded a total of 150 movement trials (75 trials per movement class) and 4 rest trials for the training paradigm. For the test paradigm, there were 4 movement trials and 1 rest trial in each of the 6 runs during session 1, and 5 runs during session 2. The study provided participants with instructions on how to perform the movements and when to report any movement attempts. The training and test paradigms for classification are given in *Fig.2* [Ofner et al., 2019].

**Figure 2:**
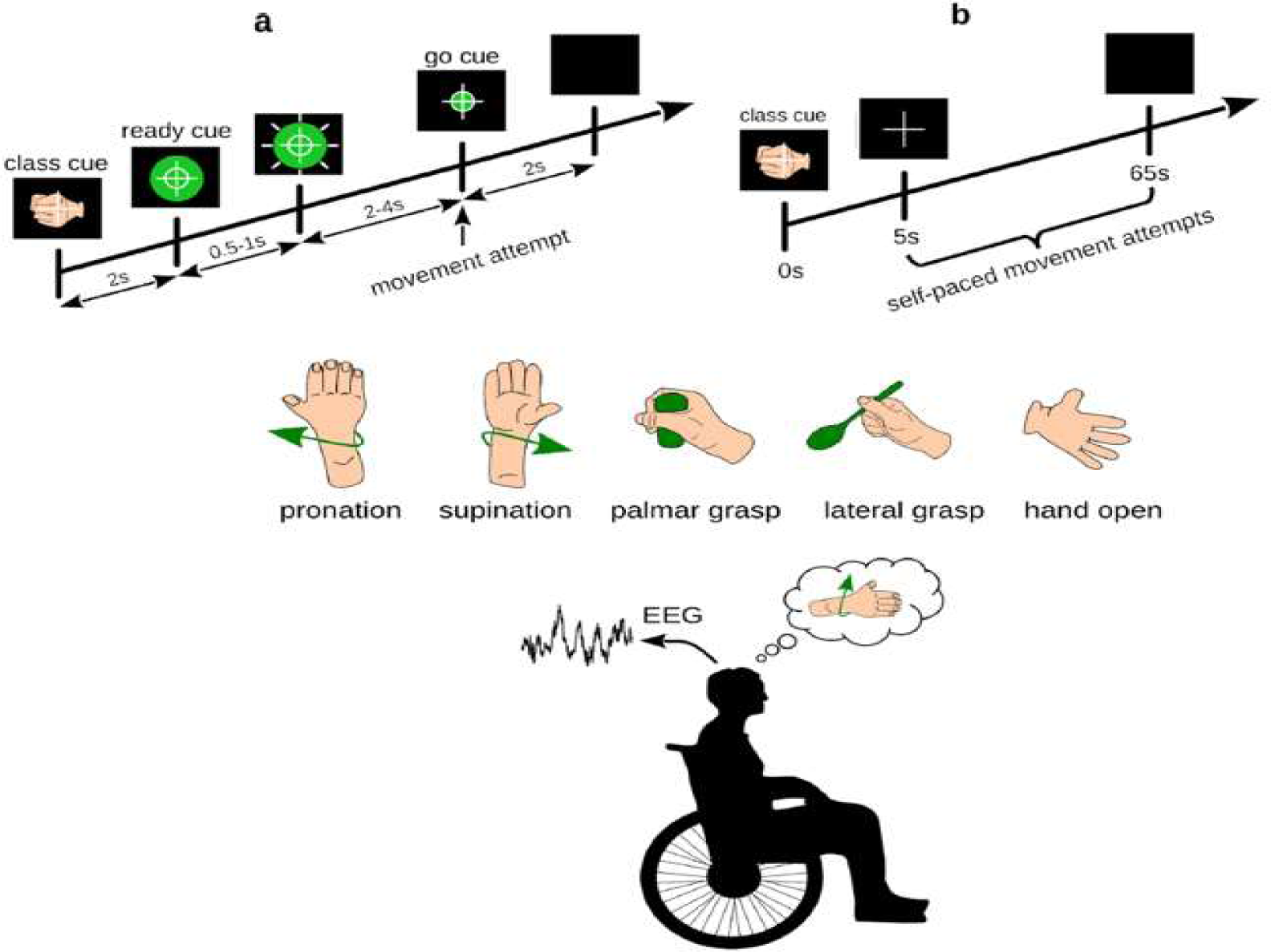
Experimental design paradigms for classification. a) training paradigm b) test paradigm [Ofner et al., 2019].

### 2.2. Data Acquisition and Preprocessing

The study utilized a 61-electrode EEG and 3-electrode EOG system to record biosignals from frontal, central, parietal, and temporal areas, as well as above the nasion and below the outer canthi of the eyes. A left earlobe reference and AFF2h ground were employed while recording signals with four 16-channel g.USBamps biosignal amplifiers and a g.GAMMAsys/g.LADYbird active electrode system, which is presented in *Fig.3*.

**Figure 3:**
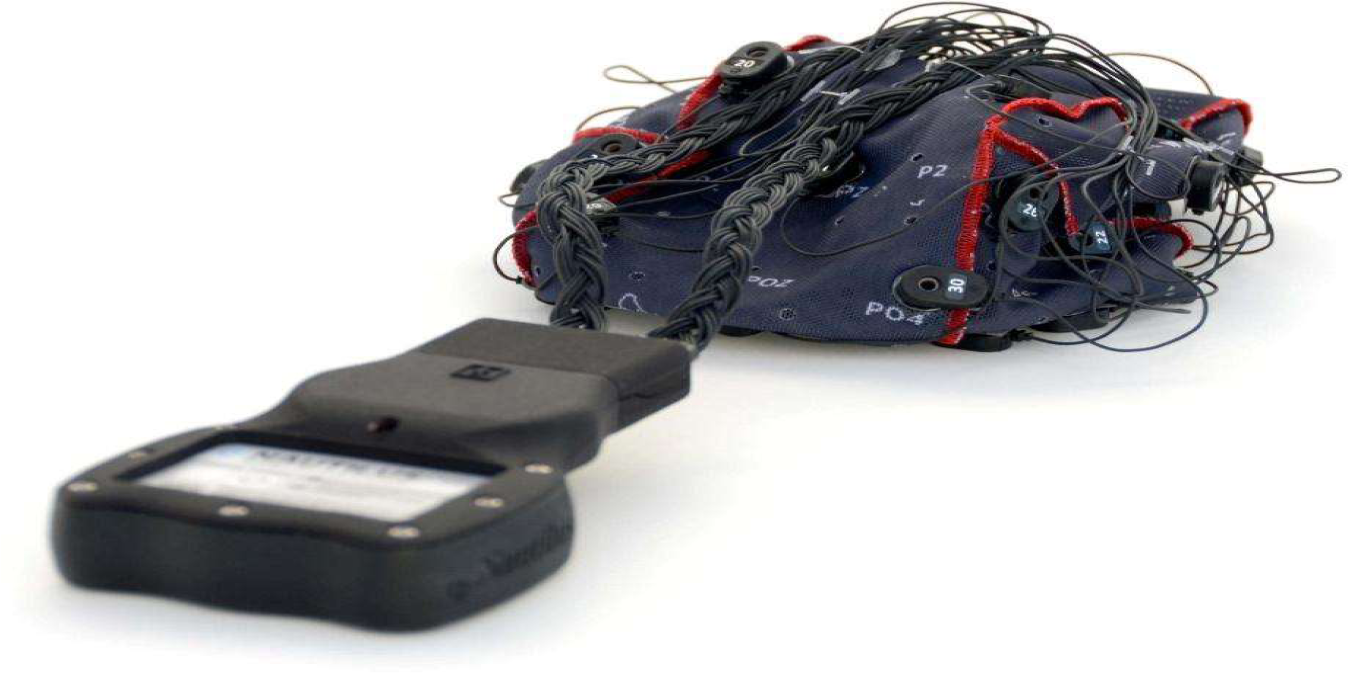
g.GAMMAsys/g.LADYbird active electrode system.

The purpose of g.GAMMAsys is to simplify and expedite the process of setting up experiments for EEG/ECG/EMG/EOG recording while ensuring excellent signal quality through the use of a comfortable cap. g.GAMMAsys is compatible with all g.tec amplifiers and offers various types of active and passive electrodes. These electrodes are connected to g.GAMMAbox, which is then connected to g.USBamp. For EEG recordings, the electrodes can be mounted onto the head using the g.GAMMAcap, while for ECG/EMG/EOG recordings, they can be placed on the body. The g.GAMMAcap can be customized with electrodes specific to a particular experiment, such as the P300 speller. Even during cleaning, the electrodes remain inside the cap, allowing for quick preparation and cleaning, thus significantly expediting the experiment process.

The signals were sampled at 256 Hz and filtered using an 8th-order Chebyshev filter with a band-pass range of 0.01 Hz to 100 Hz, and a notch filter at 50 Hz was applied to suppress power line interference. The collected data were stored in the GDF Format, with each run stored in a separate GDF file, named after the participant’s code name and run number. The data set included 15 runs per participant, consisting of 9 attempted movement runs, 3 eye movement runs, and 3 rest runs, with event codes used to encode cues and other events. The study also included an online paradigm, with two online sessions for participant P09, including training runs and test runs, which included EEG and EOG data, as well as data glove data, classifier output data, and button press data [Ofner et al., 2019]. The standard deviation was applied to the EEG signals after the filtering process. Standard deviation is a statistical measure that indicates how much the data is dispersed from the mean value. In EEG signal processing, the standard deviation is commonly used as a preprocessing step for feature extraction to reduce the dimensionality of the data and to capture relevant information about the signal. One of the main advantages of using standard deviation as a preprocessing step in EEG signal classification is that it provides a robust estimate of the variability of the signal across time. This is particularly useful for EEG signals, which are highly dynamic and subject to noise and artifacts. By computing the standard deviation of the signal, we can capture both the temporal and spectral characteristics of the EEG signal, which can be used to distinguish between different brain states or cognitive processes. The use of standard deviation as a preprocessing step in EEG signal classification has been shown to improve the accuracy and reliability of EEG-based classification models.

The formula of the standard deviation is given in *Eq.1*

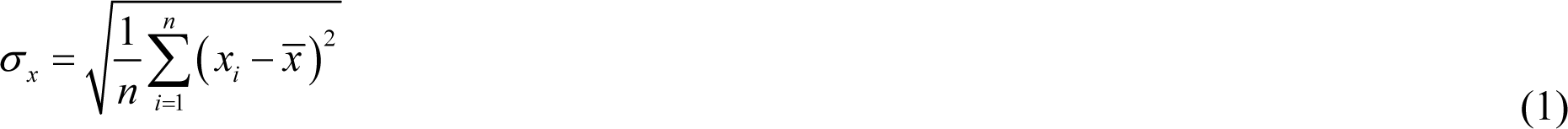

where 𝑛 is the number of samples, and *^x^* is the mean value of the samples.

### 2.3. Feature Selection Method

In the field of machine learning, feature selection is an essential process that is equally as important as feature extraction. An excessive number of features in a system can cause computational overload and increased model complexity. Hence, eliminating unused data from EEG signals before classification can significantly improve the system’s speed.

The feature selection process is crucial because it identifies the most relevant features of the signal that represent the EEG features to be classified. BCI systems often have high-dimensional feature vectors, and feature selection helps reduce the number of input variables for the classifier. Feature selection methods do not alter features but exclude certain ones based on predetermined criteria.

The goal of feature selection is to achieve the best results by processing the smallest amount of data possible. This process eliminates attributes that are irrelevant or redundant for simpler classification models, thus reducing the likelihood of overfitting in datasets with too many features or not enough observations. Feature selection methods are categorized into filter, wrapper, and embedded methods.

Embedded Feature Selection (EFS) is an efficient method for feature selection, and the random forest algorithm is a popular embedded method used to eliminate uninformative or unnecessary features. Tree-based feature selection is another method that takes advantage of the interpretable nature of the tree model. Decision trees are used as an embedded method of feature selection, where the most relevant features are selected and ranked based on their importance score.

The decision tree induction algorithm is used to learn decision tree classifiers, where each node represents an attribute test, and each branch specifies the class prediction. The information gain metric is used to select the best partitioning attribute for feature selection based on the attribute with the highest information gain.

Feature selection is a critical process that reduces the number of input variables for the classifier, eliminates irrelevant or redundant features, and reduces the likelihood of overfitting. Embedded methods, such as random forests and decision trees, are efficient and accurate feature selection methods used in machine learning [Li et.al, 2022; Bekiryazici et al., 2020; Primartha et al., 2019].

### 2.4. Deep Learning Methodology

Deep learning is a powerful machine learning technique that involves the use of multiple hidden layers to perform a series of nonlinear operations. These operations are performed using neural networks, which can learn complex functions that can distinguish between different classes of responses in classification problems.

One type of neural network that has gained considerable popularity in recent years is the recurrent neural network (RNN), which is particularly effective in extracting higher-dimensional dependencies from sequential data such as EEG time series. RNNs consist of units that have connections not only between subsequent layers but also among themselves, allowing them to receive information from previous inputs. However, traditional RNNs have difficulty learning long-term dynamics due to the disappearing and exploding gradient problems.

To address these problems, a type of RNN called Long Short-Term Memory (LSTM) was introduced. LSTMs are designed to learn both long-term and short-term dependencies, and they replace simple neurons with LSTM units, each consisting of four main components: an input gate, a neuron with self-repetitive connectivity, a forget gate, and an output gate. These components allow the LSTM to selectively recall patterns over long periods, making it particularly effective in sequence prediction and time-series prediction.

In recent years, LSTMs have gained tremendous momentum and prevalence for various applications, including classification from EEG data. LSTMs have widespread uses in real-life problems due to their ability to selectively recall patterns over long periods. The architecture of LSTMs consists of memory cells that hold the previous state and input information. These cells decide which data to keep or delete and combine the previous state with the current memory and the input data, eliminating long-term dependencies and making it possible to maintain datasets.

The forget gate in the LSTM architecture is used to reset the state and facilitate the learning of connections in the input gate. Therefore, the use of LSTMs in classification problems can be highly effective, particularly when dealing with complex sequential data such as EEG time series [Zhang et al., 2020; Agarwal et al., 2020; Tayeb et al., 2019].

In recent years, Convolutional Neural Networks (CNNs) have emerged as a powerful tool for addressing complex tasks across various domains. These networks are inspired by the hierarchical organization of processing units in the animal visual system. CNNs are designed to automatically learn features from raw input data without manual intervention, making them highly effective for processing different data types, including spectrograms.

CNNs are composed of stacked convolutional layers that employ convolutional filters to scan the input layer nodes and calculate the output. Each layer in the network utilizes different convolutional filters, and the network learns the appropriate filters when exposed to training data. CNN architectures encompass several layers such as the Convolution Layer, Batch Normalization (BN) Layer, Activation Layer, Pooling Layer, Dropout Layer, Flatten Layer, and Fully Connected (FC) Layer.

The Convolution Layer is essential in CNNs as it enables feature detection in the data. The filter matrix dimensions are typically 3×3, 5×5, or 7×7. The BN Layer is employed to address issues like gradient loss and minimal learning by standardizing input values. The Activation Layer applies non-linear functions (e.g., sigmoid, tanh, ReLU) to determine the output. The Pooling Layer, similar to the Convolution Layer, reduces the computational burden by decreasing the image size. The Dropout Layer prevents overfitting by randomly deactivating a portion of neurons during training. The Flatten Layer transforms data into a one-dimensional array, preparing it for the FC Layer. In the FC Layer, each input connects with all neurons.

Normalization is a crucial preprocessing step for the RNN component of the model to mitigate negative effects caused by outliers in the dataset. Data normalization aims to eliminate redundancy and inconsistency in the database, controlling neural network complexity and achieving generalizable performance across various applications. Batch Normalization (BN) is the most commonly used normalization method. BN normalizes the input layer through re-centering and re-scaling, making artificial neural networks faster and more stable. BN accelerates the training speed of deep neural networks by reducing the internal covariate shift, which refers to the change in the distributions of internal nodes in a deep network. BN also acts as a regulator, potentially replacing the need for dropout and preventing overfitting [Peng, 2018; Awais et al., 2021; Baumgartner et al., 2017; Poernomo&Kang, 2018; Bengio et al., 2007].

Adaptive Momentum (Adam) optimizer, which has gained widespread adoption in deep learning due to its efficiency and effectiveness, has been used. Adam is a stochastic optimization method that only requires first-order gradients and has low memory requirements. It computes adaptive learning rates for individual parameters using estimates of the first and second moments of the gradients, hence the name adaptive moment estimation.

A major advantage of Adam is its combination of two other popular optimization methods, AdaGrad and RMSProp. AdaGrad performs well with sparse gradients, while RMSProp is effective in online and non-stationary environments. Adam’s combination allows it to handle sparse gradients in noisy datasets and naturally perform gradual size annealing. Additionally, it can efficiently handle large datasets with minimal computational overhead.

However, the Adam optimizer has certain limitations. It may not always converge to the optimal solution and can encounter weight loss problems. Moreover, recent optimization algorithms have proven faster and more effective in specific scenarios. Nonetheless, the default hyperparameter values of Adam are generally sufficient for most problems, and it remains a popular and reliable optimization method in the field of deep learning [Barakat&Bianchi, 2021; Krizhevsky et al., 2017; Lecun et al., 1998].

## 3. Results

After performing the standard scaling process, the data has a size of [64,3775000]. It comprises five classes that are equally distributed. Given that the EEG device’s sampling frequency is 250 Hz, the data is reshaped to [15100,250,64]. When feeding the data matrix into the classifier algorithm, it follows a specific order. The rows represent the data package (250) at a particular sample time, while the columns represent the EEG and ECoG channels (64). The samples are organized in the 3rd dimension with a count of 15100. To label the data, the one-hot encoder method was employed, resulting in labels with a shape of [15100, 5]. Following the reshaping and labeling steps, the data is further processed by shuffling it and splitting it into train, test, and validation datasets. The dimensions of the train, test, and validation data are presented in *Table 1*.

**Table 1:** The size of the train, test, and validation data.

| **Data Type** | **Size** |
| --- | --- |
| **Train Data** | [11325, 250, 64] |
| **Test Data** | [1510, 250, 64] |
| **Validation Data** | [2265, 250, 64] |
| **Train Label** | [11325, 5] |
| **Test Label** | [1510, 5] |
| **Validation Label** | [2265, 5] |

### 3.1. 1D CNN+LSTM model

In this study, a 1D CNN and LSTM model is used to create a combined system, which aims to improve the accuracy of the model in predicting sequences.

The model consists of multiple layers that are designed to process the input data in a way that captures the essential features of the sequence. The input data, in this case, is a 3D tensor with shape (timesteps, dims), where timesteps refer to the length of the sequence and dims refer to the number of features in each timestep. The model architecture can be broken down into the following steps:

The model architecture we will be discussing is a combination of Convolutional Neural Networks (CNNs) and Long Short-Term Memory (LSTM) networks. The model is built using the TensorFlow Keras Sequential API, which allows us to stack layers on top of each other in a linear fashion and provides a high-level interface for building deep learning models.

The first layer of the model is a 1D convolutional layer with 24 filters and a kernel size of 8. This layer takes input in the shape of (timesteps, and dims), where timesteps are the number of time steps in the input sequence and dims are the number of dimensions in each time step. The activation function used in this layer is ReLU (Rectified Linear Unit), which is a popular choice in deep learning for its simplicity and effectiveness in solving the vanishing gradient problem. The next layer is a max pooling layer with a pool size of 4, which reduces the dimensionality of the output from the previous layer. The second convolutional layer has 8 filters and a kernel size of 4. It also uses ReLU as the activation function and has a stride of 2, which reduces the output dimensionality further. The next layer is an LSTM layer with 45 units, which takes input from the previous layer and returns a sequence of outputs. The activation function used in this layer is tanh, which is a common choice for LSTM networks. Additionally, this layer has a dropout rate of 0.2, which helps prevent overfitting. The second LSTM layer has 25 units and takes input from the previous LSTM layer. This layer does not return a sequence of outputs, but instead returns a single output. After the LSTM layers, we add a dropout layer with a rate of 0.5, which randomly drops out 50% of the neurons to reduce overfitting. The final layer is a dense layer with 5 output units, which uses softmax activation to predict the class probabilities. This layer also has weight decay regularization applied to it, with a regularization parameter of 0.001.

The model is compiled using the categorical cross-entropy loss function and the Adam optimizer, which is a popular choice in deep learning for its effectiveness in handling noisy data and convergence speed. The model is trained using early stopping, which stops the training process if the validation loss does not improve for a certain number of epochs.

The model is trained for 150 epochs with a batch size of 128. The model is evaluated on test data, and the test accuracy is reported.

The use of both CNN and LSTM layers enables the model to capture both spatial and temporal features of the sequence. Additionally, the use of regularization techniques such as dropout and weight decay helps prevent overfitting and improves the generalization ability of the model. The architecture of the model is given in *Fig.4*.

**Figure 4:**
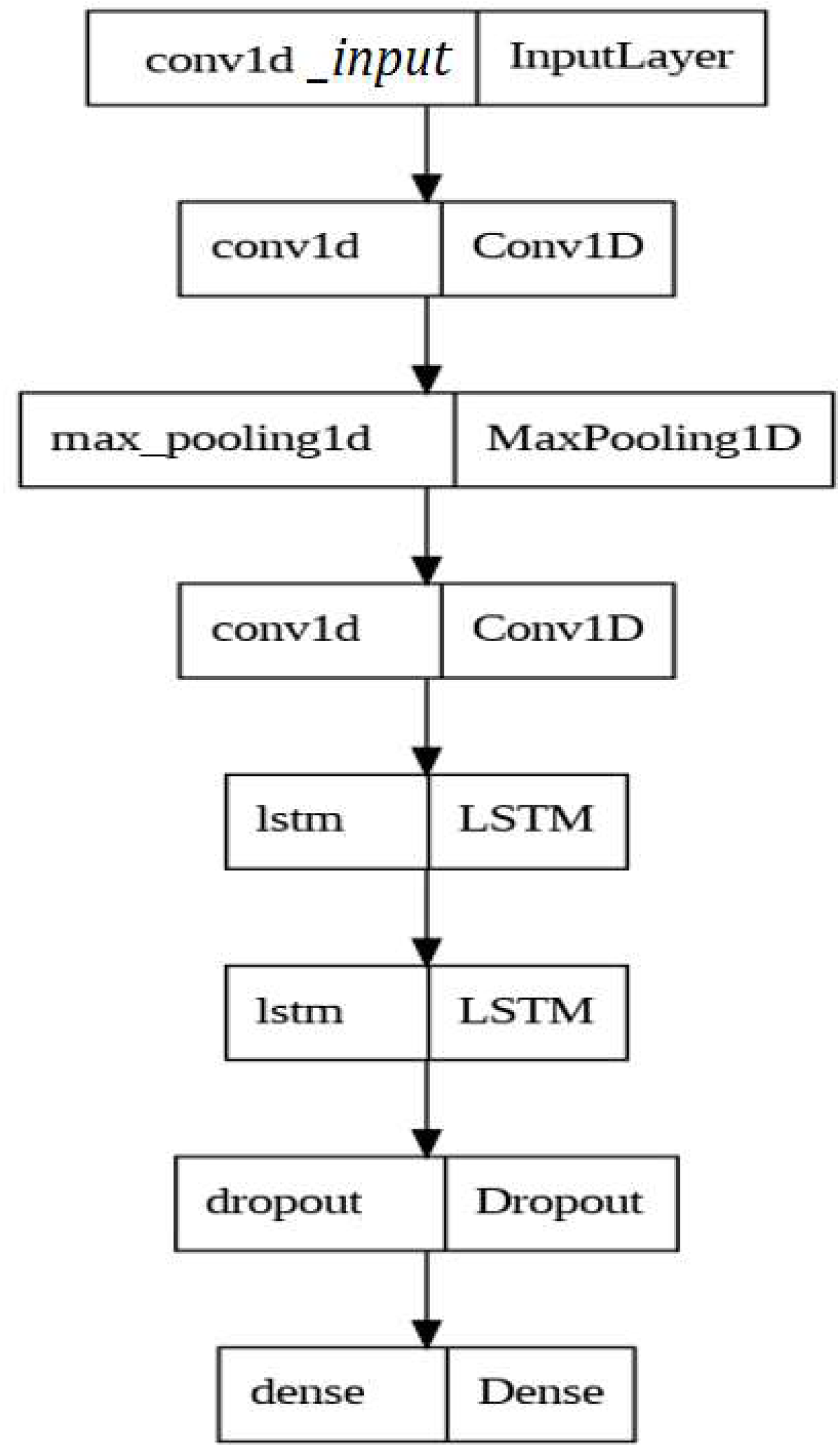
The architecture of the proposed deep learning model.

Training and test model accuracy and loss function values for the first 150 epochs are given in *Fig.5a* and *Fig.5b*, respectively.

**Figure 5:**
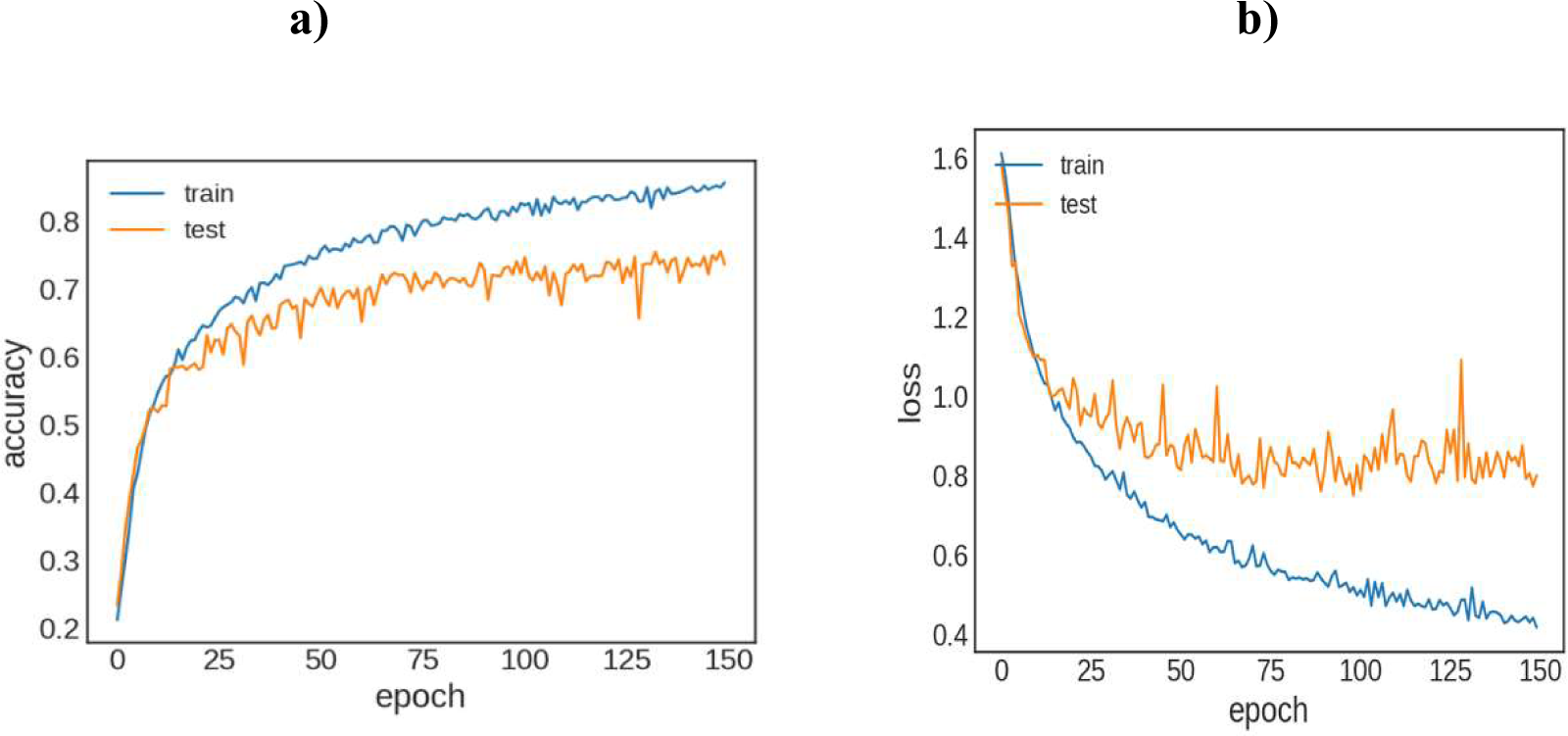
a) Training and test model accuracy, b) Training and test loss function values.

The confusion matrix is given in *Table 2*.

**Table 2:**
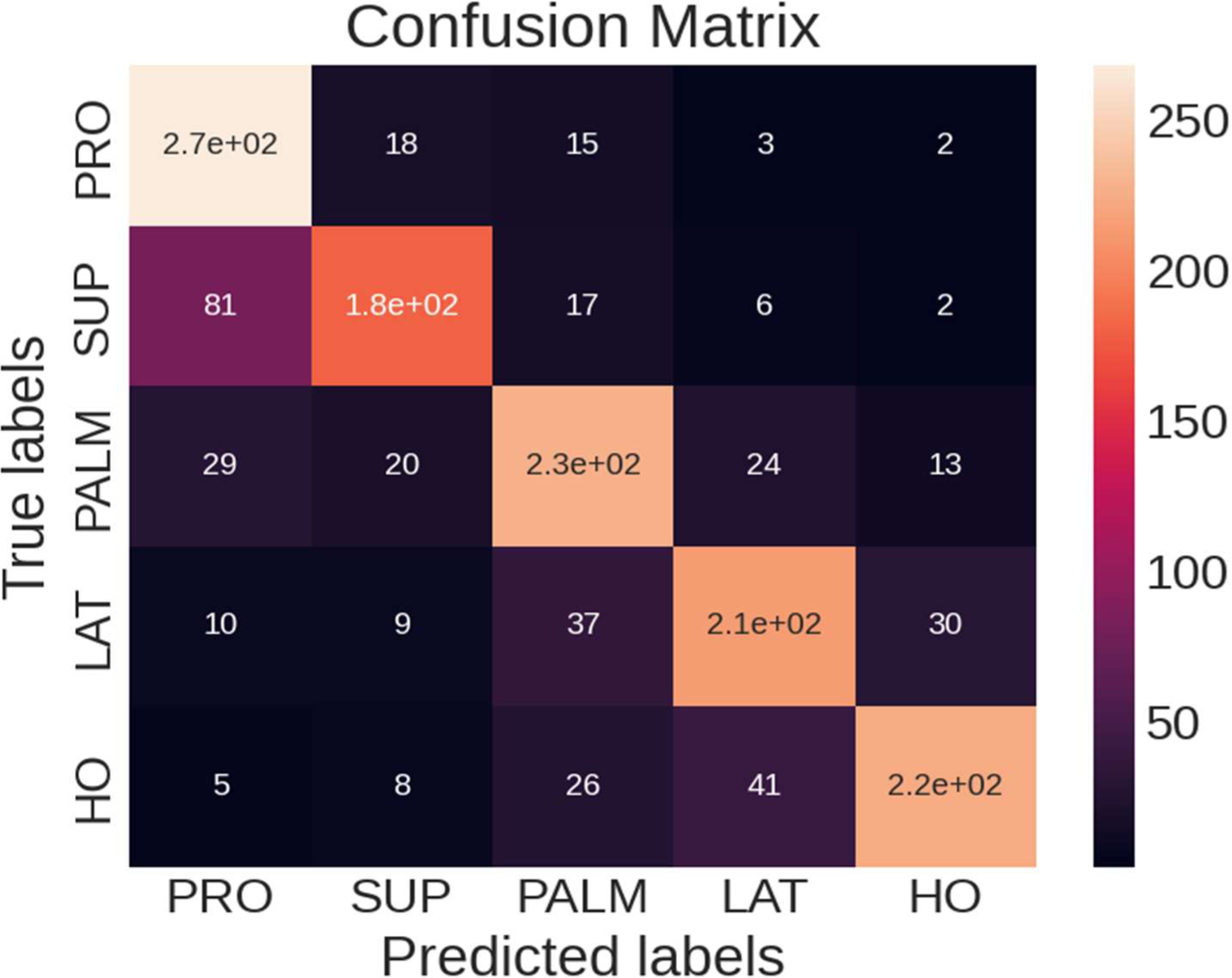
The confusion matrix of the model.

The model’s test accuracy is reported as 73.775%. This metric represents the proportion of correctly classified instances in the test dataset.

We also assess the model’s performance using the Kappa value, which is calculated to be 0.63. Kappa evaluates the agreement between predicted and actual values, taking into account the agreement occurring by chance.

In terms of the model’s overall effectiveness, the mean f1 score is measured as 0.738. This score combines precision and recall, providing an assessment of the model’s ability to balance accurate predictions and complete coverage of relevant instances.

Specifically, the mean precision score is determined as 0.746, reflecting the model’s ability to accurately identify positive instances among the predicted positives.

Additionally, the mean recall score is calculated as 0.738, indicating the model’s capacity to capture a significant portion of actual positive instances among all positive cases in the dataset. By considering these metrics together, we can better understand and evaluate the performance of the model in terms of accuracy, agreement, precision, and recall.

### 3.2. 1D CNN+LSTM model with Feature Selection

Building a machine learning system poses challenges when dealing with high-dimensional and noisy data. One effective technique to address this is feature selection, which aims to reduce the dimensionality of the data by identifying the most important features that contribute significantly to the system’s predictive performance. In this article, we compare three feature selection algorithms: tree-based embedded random forest, wrapper-based recursive feature elimination, and filter-based chi-square feature selection. The goal is to reduce the number of electrodes from 64 to improve the system’s efficiency. The first method, tree-based embedded random forest, estimates feature importance using decision trees. By fitting a random forest model to the data, it utilizes the mean decrease impurity metric to determine feature importance and selects the most crucial features accordingly. The second method, wrapper-based recursive feature elimination, adopts a model-based approach for feature selection. It recursively removes features from the dataset, fits a model to the remaining features, evaluates model performance, and eliminates the feature with the least contribution. This process continues until the desired number of features is achieved. The third method, filter-based chi-square feature selection, leverages statistical tests to select informative features. By calculating the chi-square statistic for each feature, it identifies and selects the features with the highest statistics. After applying these methods to the dataset, the most important features are selected using the Xgboost method, a widely-used gradient-boosting algorithm for feature selection. Next, the selected features are fed into a 1D CNN+LSTM classifier, a deep-learning model suitable for sequential data analysis. This classifier consists of a convolutional layer for extracting local features and an LSTM layer for capturing temporal dependencies between these extracted features. In summary, feature selection plays a crucial role in reducing the dimensionality of high-dimensional and noisy data. In this article, we examined and compared tree-based embedded random forest, wrapper-based recursive feature elimination, and filter-based chi-square feature selection algorithms. We also explored the Xgboost method for assessing feature importance and the 1D CNN+LSTM classifier for sequential data analysis. Applying these methods improved system performance by selecting the most relevant features. It’s important to note that selecting too few features may result in information loss while selecting too many features can lead to overfitting and decreased performance. To optimize the number of selected features, various methods can be employed. In our case, cross-validation was used to estimate model performance with different feature subsets. The dataset was divided into training and validation sets, and the feature selection method was applied to the training set. The selected features were then used for training the model, and its performance was evaluated on the validation set. This process was repeated with different numbers of selected features, and the number of features yielding the best performance on the validation set was chosen. Wrapper-based recursive feature elimination algorithms have built-in mechanisms to optimize the number of selected features. These algorithms utilize cross-validation and iterative methods to determine the optimal number of features for a given problem. After optimization, it was found that the best classification accuracy was achieved with a subset of 32 selected features. The locations of the selected features are given in *Fig.6*.

**Figure 6:**
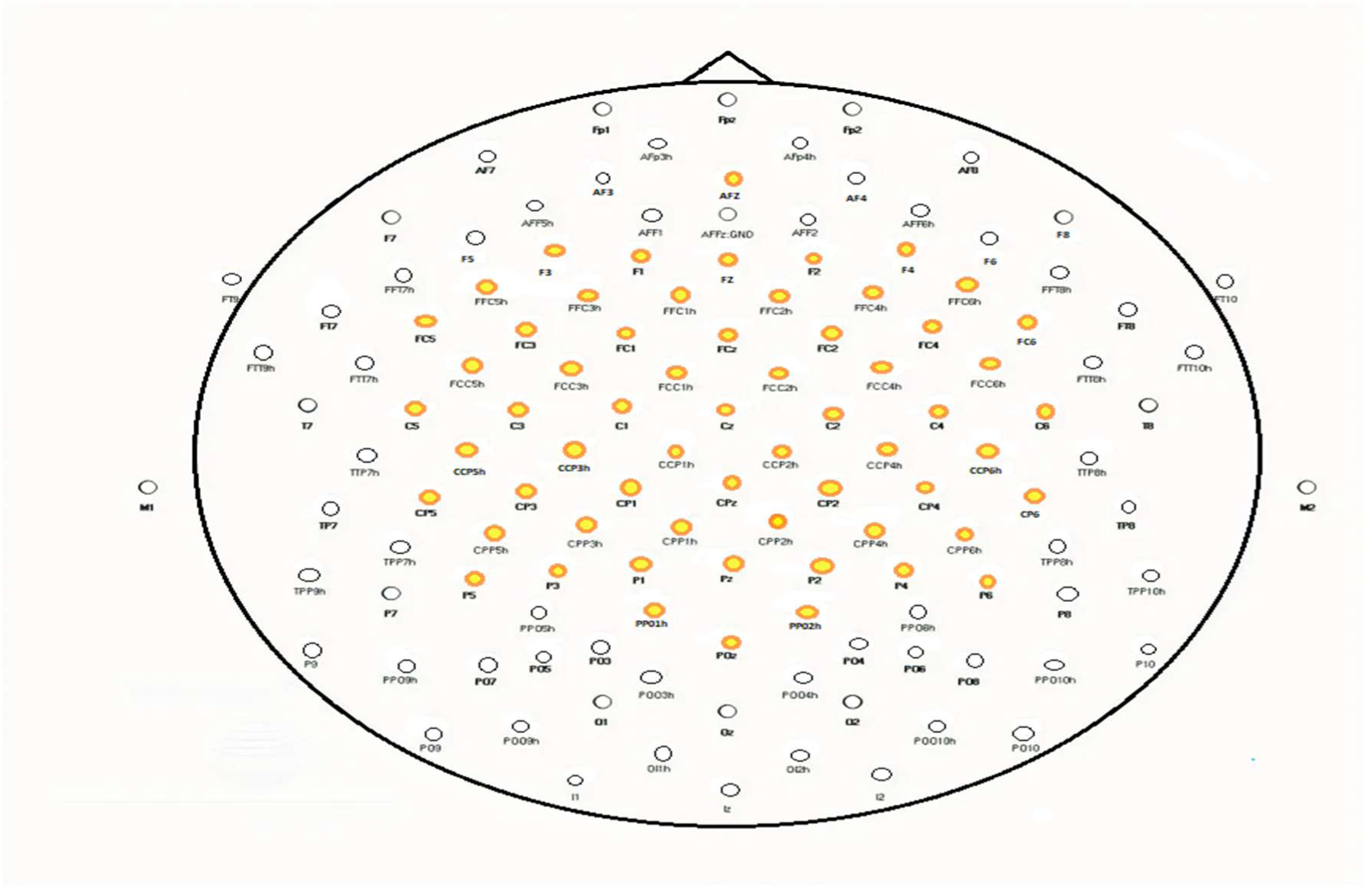
Location and nomenclature of a 64-channel cap according to the 5% electrode placement scheme: frontal pole (Fp), antero-frontal (AF), frontal (F), central (C), parietal (P), temporal (T), occipital (O), and mastoid (M). The numbering starts at the midline (z for zero) increasing with distance. Even numbers indicate right-sided placement with odd numbers on the left. [Martin et al., 2014].

The labels and the corresponding numbers of electrodes that are mapped to the EEG cap are given: [’AFz’]→ 1, [’F3 ’] → 2, [’F1’] → 3, [’Fz’] → 4, [’F2’] → 5, [’F4’] → 6, [’FFC5h’] → 7, [’FFC3h’] → 8, [’FFC1h’] → 9, [’FFC2h’] → 10, [’FFC4h’] → 11, [’FFC6h’] → 12, [’FC5’] → 13, [’FC3’] → 14, [’FC1’] → 15, [’FCz’] → 16, [’FC2’] → 17, [’FC4’] → 18, [’FC6’] → 19, [’FCC5h’] → 20, [’FCC3h’] → 21, [’FCC1h’] → 22, [’FCC2h’] → 23, [’FCC4h’] → 24, [’FCC6h’] → 25, [’C5’] → 26, [’C3’] → 27, [’C1’] → 28, [’Cz’] → 29, [’C2’] → 30, [’C4’] → 31, [’C6’] → 32, [’CCP5h ’] → 33, [’CCP3h’] → 34, [’CCP1h’] → 35, [’CCP2h’] → 36, [’CCP4h’] → 37, [’CCP6h’] → 38, [’CP5’] → 39, [’CP3’] → 40, [’CP1’] → 41, [’CPz’] → 42, [’CP2’] → 43, [’CP4’] → 44, [’CP6’] → 45, [’CPP5h’] → 46, [’CPP3h’] → 47, [’CPP1h’] → 48, [’CPP2h’] → 49, [’CPP4h’] → 50, [’CPP6h’] → 51, [’P5’] → 52, [’P3’] → 53, [’P1’] → 54, [’Pz’] → 55, [’P2’] → 56, [’P4’] → 57, [’P6’] → 58, [’PPO1h’] → 59, [’PPO2h’] → 60, [’POz’] → 61, [’eog-l ’] → 62, [’eog-m’] → 63, [’eog-r’] → 64 The selected features obtained from three different methods are listed in *Table 3*.

**Table 3:** The features obtained from the feature selection methods.

|  | Feature Selection Method |  |  |
| --- | --- | --- | --- |
|  | Filter-based<br>(Chi-Squared) | Embedded<br>(Random Forest) | Wrapper-based<br>(RFE) |
| **Selected**<br>**Features** | 4 | 1 | 1 |
|  | 5 | 2 | 3 |
|  | 8 | 3 | 4 |
|  | 12 | 6 | 5 |
|  | 14 | 7 | 8 |
|  | 15 | 10 | 9 |
|  | 16 | 11 | 11 |
|  | 17 | 12 | 12 |
|  | 18 | 13 | 14 |
|  | 19 | 14 | 17 |
|  | 20 | 15 | 18 |
|  | 21 | 17 | 21 |
|  | 22 | 18 | 22 |
|  | 27 | 20 | 28 |
|  | 29 | 21 | 30 |
|  | 31 | 24 | 31 |
|  | 33 | 28 | 41 |
|  | 37 | 29 | 42 |
|  | 38 | 30 | 43 |
|  | 41 | 32 | 44 |
|  | 42 | 36 | 47 |
|  | 43 | 42 | 50 |
|  | 44 | 43 | 51 |
|  | 45 | 45 | 52 |
|  | 50 | 48 | 55 |
|  | 52 | 52 | 56 |
|  | 55 | 54 | 57 |
|  | 59 | 58 | 58 |
|  | 60 | 59 | 60 |
|  | 62 | 60 | 62 |
|  | 63 | 62 | 63 |
|  | 64 | 64 | 64 |

In this study, three different feature selection methods were employed to select features for analysis. The selected features from each method were then used as inputs for training a 1D CNN + LSTM deep learning model, allowing for comparative analysis.

A total of 32 features were selected by each method, and these features were used as inputs for training the model. The training process was repeated six times for each method. The resulting test accuracy values were used to calculate p-values, which measure the statistical significance of the differences between the methods.

The p-value between the Random Forest (RF) and Recursive Feature Elimination (RFE) methods was found to be P = 0.0000122, indicating a statistically significant difference between these two methods. Similarly, the p-value between RF and Chi-square methods was calculated as P = 0.00000746, also suggesting a significant difference between RF and Chi-square.

On the other hand, the p-value between RFE and Chi-square methods was determined to be P = 0.78321757709, which is greater than the commonly used significance level of 0.05. This implies that there is no statistically significant difference in performance between RFE and Chi-square.

The test accuracy values obtained after six training runs for each method were recorded. For RF, the test accuracy values were [0.80244, 0.81674, 0.82006, 0.81912, 0.80331, 0.81325], for RFE they were [0.77351, 0.78300, 0.77921, 0.77145, 0.78001, 0.78940], and for Chi-square they were [0.76995, 0.78117, 0.77154, 0.78166, 0.78132, 0.78477].

The comparison in terms of test and train accuracy is given in *Table 4*.

**Table 4:** Test and train accuracies of the models subjected to different feature selection methods.

| Model | Test Accuracy (%) | Train Accuracy (%) |
| --- | --- | --- |
| **RF+1D CNN+LSTM** | 81.248 | 82.934 |
| **RFE+1D CNN+LSTM** | 77.943 | 81.737 |
| **Chi-square+ 1D CNN+LSTM** | 77.840 | 81.332 |

Based on the p-values and the results in the table, it can be concluded that the Random Forest method (RF) yielded the best performance among the three feature selection methods. Therefore, the features selected by the Random Forest method were chosen for further analysis and model training.

The confusion matrix is given in *Table 5*.

**Table 5:**
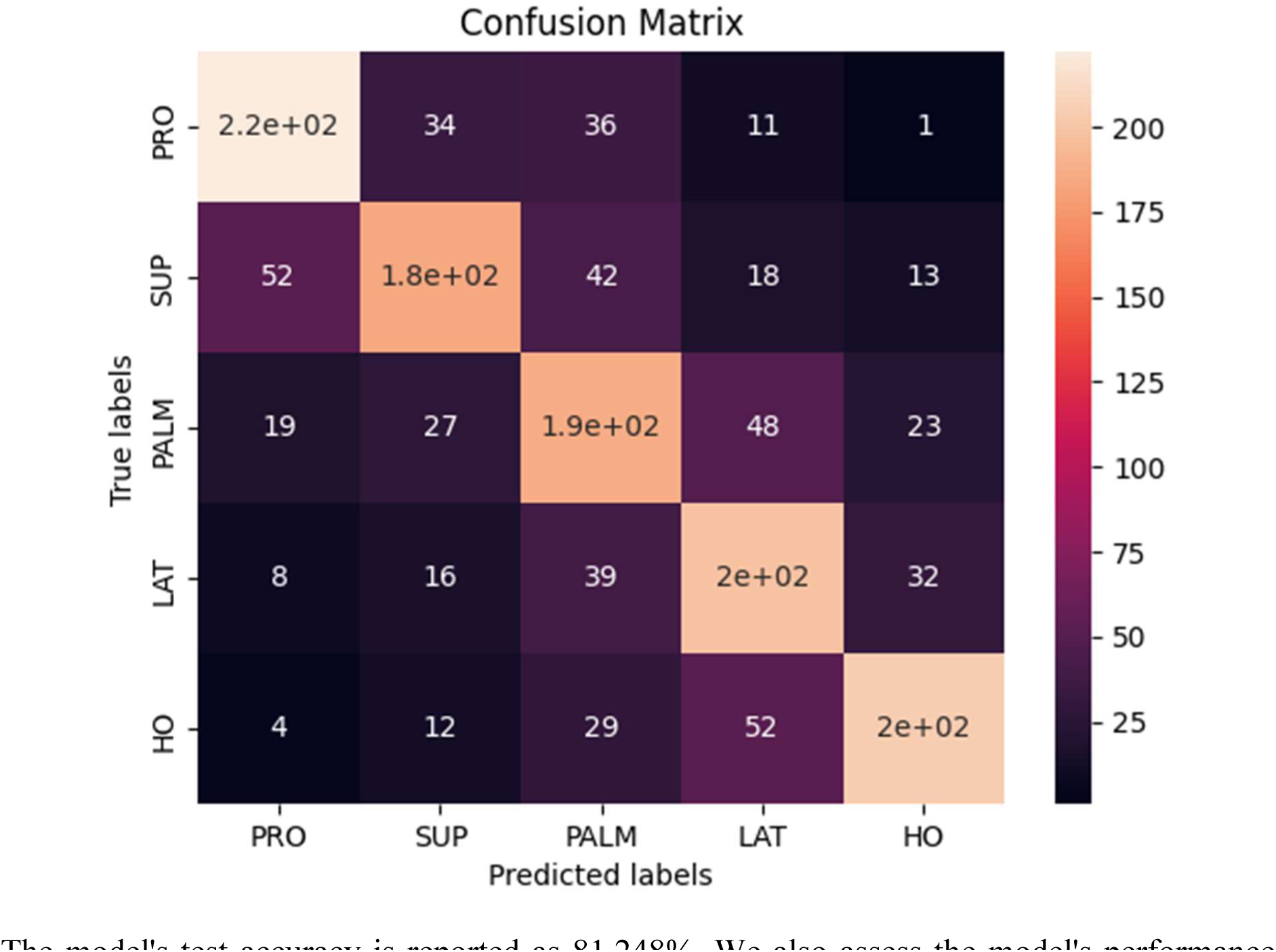
The confusion matrix of the model (Features selected with the RF method).

The model’s test accuracy is reported as 81.248%. We also assess the model’s performance using the Kappa value, which is calculated to be 0.648. In terms of the model’s overall effectiveness, the mean f1 score is measured as 0.81. Specifically, the mean precision score is determined as 0.844. Additionally, the mean recall score is calculated as 0.779.

Using the random forest feature selection method, we identified and measured the weights of 32 electrodes.

To determine the most influential electrodes, we applied the XGBoost (eXtreme Gradient Boosting) algorithm, specifically the Feature Importance functionality. By analyzing the results, we found that electrodes 28, 36, 43, 48, and 59 exhibited XGBoost values higher than 0.02, placing them in the top row of *Table 6*.

**Table 6:** The scores of the selected features obtained from the feature importance analysis.

| Selected Feature | XGBoos t Score | Selected Feature | XGBoos t Score | Selected Feature | XGBoos t Score | Selected Feature | XGBoos t Score |
| --- | --- | --- | --- | --- | --- | --- | --- |
| 1 | 0.01935 | 13 | 0.01442 | 28 | 0.02945 | 48 | 0.02057 |
| 2 | 0.01613 | 14 | 0.01979 | 29 | 0.01662 | 52 | 0.01627 |
| 3 | 0.01591 | 15 | 0.01662 | 30 | 0.01675 | 54 | 0.01259 |
| 6 | 0.01362 | 17 | 0.01662 | 32 | 0.01329 | 58 | 0.01487 |
| 7 | 0.01366 | 18 | 0.01870 | 36 | 0.02050 | 59 | 0.02027 |
| 10 | 0.01914 | 20 | 0.01869 | 42 | 0.01770 | 60 | 0.01609 |
| 11 | 0.01865 | 21 | 0.01700 | 43 | 0.02515 | 62 | 0.01595 |
| 12 | 0.01743 | 24 | 0.01596 | 45 | 0.01415 | 64 | 0.01495 |

These electrodes have been identified as having significant importance in the dataset based on their weights and XGBoost scores.

XGBoost is a powerful machine learning algorithm that utilizes gradient boosting techniques to create an ensemble of weak prediction models, typically decision trees, and combines their predictions to make more accurate predictions. In addition to its strong predictive capabilities, XGBoost provides a useful feature importance analysis, which helps in understanding the relative importance of different features in the dataset for making predictions.

Feature importance in XGBoost is determined by considering the contribution of each feature to the reduction of the loss function (e.g., mean squared error) during the training process. The algorithm calculates the total gain, also known as the total reduction in the loss function, achieved by using a specific feature to make splits in the decision trees. Features that result in higher gain are considered more important.

The feature importance in XGBoost can be assessed using various techniques. One commonly used method is the “weight” metric, which represents the number of times a feature is used to make splits across all decision trees. Features with higher weights are generally considered more important.

Another method is the “cover” metric, which represents the average coverage (i.e., the number of samples affected) of a feature across all decision trees. Features that cover a larger portion of the dataset are considered more important.

Finally, the “gain” metric, as mentioned earlier, represents the average gain achieved by using a feature for making splits across all decision trees. Features with higher average gains are considered more important.

By analyzing the feature importance scores provided by XGBoost, data scientists can identify the most influential features in a dataset, gain insights into the relationships between features and the target variable, and potentially improve the model’s performance by selecting or engineering more relevant features [Wang et al., 2020; Friedmann, 2001; Raihan et al., 2023; Machado et al., 2019].

The scores for each electrode obtained from the XGBoost analysis are represented in *Fig. 7*.

**Figure 7:**
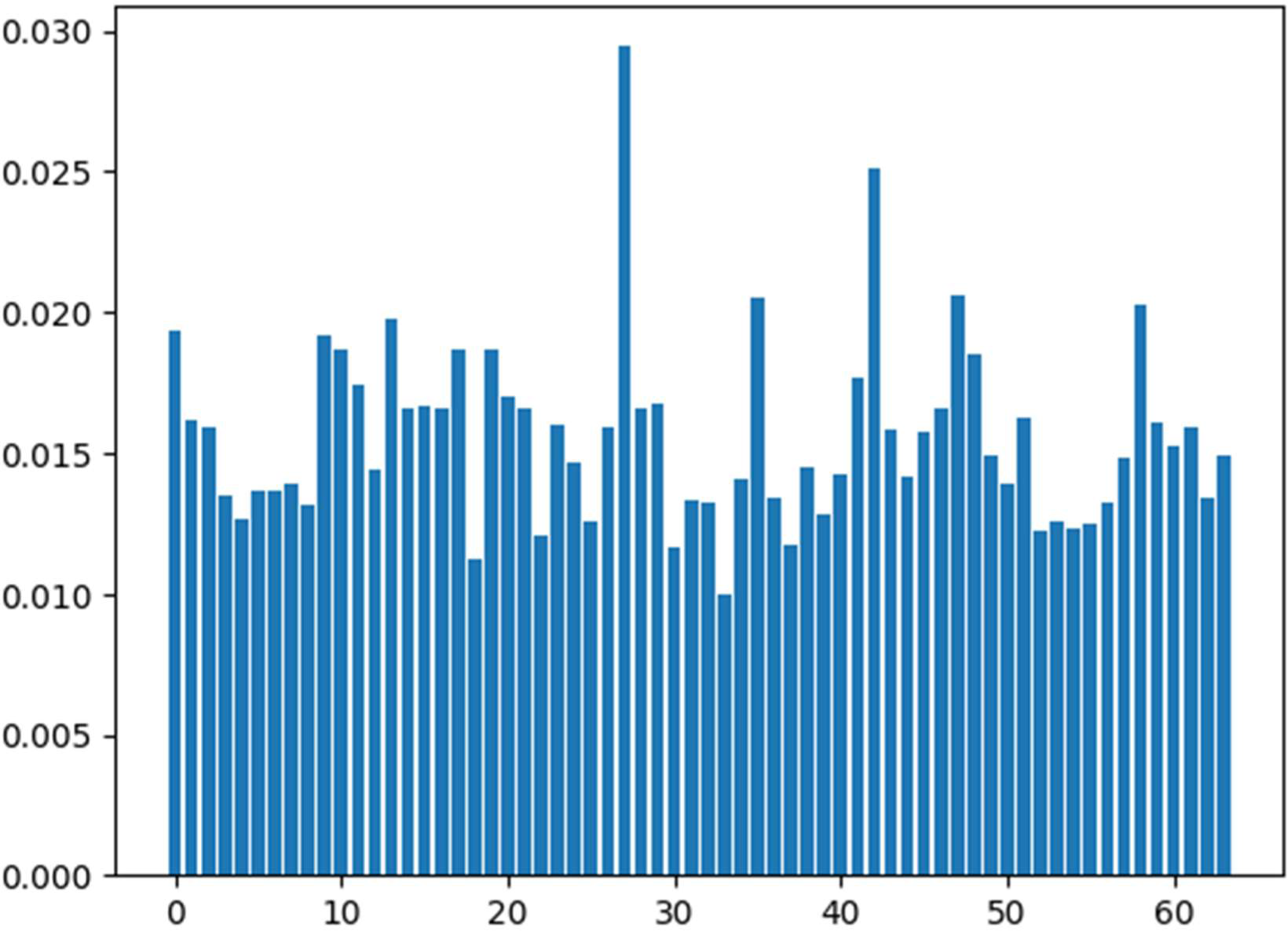
The feature importance results obtained from the XGBoost method.

### 3.3. The result after applying 10-fold cross-validation

Cross-validation is a widely used technique in data science for evaluating the performance of machine learning models and estimating their generalization ability. Specifically, 10-fold cross-validation is a common approach that divides the dataset into ten equal-sized subsets or “folds.”

The 10-fold cross-validation process can be summarized as follows:

The dataset is randomly shuffled to ensure an unbiased representation of the data across the folds.

The data is divided into ten equal-sized subsets, each containing an approximately equal distribution of samples. One subset is set aside as the “validation set” or “holdout set,” while the remaining nine subsets are used for training.

The model is trained on the training set, using nine subsets of data, and evaluated on the holdout set. This process is repeated ten times, with each of the ten subsets used as the holdout set once. At the end of the ten iterations, the evaluation metric (e.g., accuracy, precision, or mean squared error) is recorded for each fold. The evaluation metric from each fold is then averaged to provide an overall assessment of the model’s performance.

The benefits of using 10-fold cross-validation include:

More robust performance estimation: By repeating the training and evaluation process ten times with different holdout sets, we obtain a more reliable estimate of the model’s performance, reducing the impact of variability in the data.

Efficient use of data: Cross-validation allows us to utilize a larger portion of the data for both training and evaluation, compared to a simple train-test split.

Model selection and hyperparameter tuning: Cross-validation aids in selecting the best model or tuning hyperparameters. By comparing the performance of different models or hyperparameter settings across the ten folds, we can identify the most effective choices.

Detecting overfitting: Cross-validation helps in assessing whether the model is overfitting the training data. If the model performs significantly worse on the holdout sets compared to the training sets, it suggests overfitting, indicating a need for adjustments [Varma&Simon, 2006; Stone, 2006; Jiang&Wang, 2017].

We applied the 10-Fold CV to the features selected 1D CNN+LSTM model. The results are given in *Table 7*.

**Table 7:** 10-Fold cross-validation results.

| **10-Fold Cross Validation** | **Accuracy** |
| --- | --- |
| **Iteration 1** | 76.49% |
| **Iteration 2** | 75.70% |
| **Iteration 3** | 74.17% |
| **Iteration 4** | 75.76% |
| **Iteration 5** | 76.23% |
| **Iteration 6** | 75.17% |
| **Iteration 7** | 76.09% |
| **Iteration 8** | 76.23% |
| **Iteration 9** | 76.69% |
| **Iteration 10** | 74.97% |

|  | **Mean Accuracy** | **Standard Deviation** |
| --- | --- | --- |
| **10-Fold Cross Validation** | 75.75% | +/- 0.74% |

## 4. Discussion and Conclusive Summary

The research conducted aimed to enhance the classification performance of specific movements in patients with cervical spinal cord injury (SCI) using electroencephalography (EEG) data. By combining convolutional neural network (CNN) and long-short-term memory (LSTM) architectures, the study successfully revealed strong neural correlations between temporal and spatial aspects of the EEG signals associated with attempted arm and hand movements.

To improve the model’s performance, feature selection methods were applied. The Random Forest (RF) feature selection method yielded a significant improvement, resulting in a test accuracy of 81.248% when combined with the 1D CNN+LSTM model. This indicates the effectiveness of RF in selecting relevant features for classification.

Additionally, the study assessed the robustness of the proposed model against data variations using 10-fold cross-validation (CV). The mean test accuracy achieved with RF feature selection, combined with 1D CNN+LSTM and 10-fold CV, was 75.75%. Although slightly lower than the RF-enhanced model without cross-validation, this result indicates that the model maintains reasonable performance across different folds and provides a more reliable estimate of its generalization ability.

In conclusion, the research successfully contributes to the field of neuro-technology by developing EEG-controlled assistive devices through a generalized brain-computer interface (BCI) and deep learning (DL) framework. By capturing high-level spatiotemporal features and latent dependencies, the proposed model demonstrates improved classification performance for specific movements in SCI patients. The findings highlight the potential for utilizing EEG-based technologies in assisting individuals with motor impairments and provide a foundation for further advancements in the field of neuro-technology.

**Patient-informed consent:** There is no need for patient-informed consent.

**Ethics committee approval:** There is no need for ethics committee approval.

**Conflict of interest:** There is no conflict of interest to declare.

**Financial support and sponsorship:** No funding was received.

**Author contribution subject and rate:** STZM (80%), CU(20%): Design the research, data collection, and analyses and wrote the whole manuscript. STZM (30%), CU (70%): Organized the research and supervised the article write-up. STZM(15%), CU (85%): Contributed with comments on research design and slide interpretation. STZM(40%), CU (60%): Contributed with comments on manuscript organization and write-up.

## Data Availability

All data produced are available online at http://bnci-horizon-2020.eu/database/data-sets and from Zenodo at https://doi.org/10.5281/zenodo.2222268.

## References

1. Agarwal, T., Raturi, S., Vybhav, T. K., & Singh, M. (2020). Classification of EEG signal using lstms under audiovisual stimuli. 2020 International Conference on Communication and Signal Processing (ICCSP). https://doi.org/10.1109/iccsp48568.2020.9182092

2. Akmal, M. (2022). Tensor factorization and attention-based CNN-LSTM deep-learning architecture for improved classification of missing physiological sensors data. IEEE Sensors Journal, 1–1. https://doi.org/10.1109/jsen.2022.3223338

3. Awais, M., Bin Iqbal, Md. T., & Bae, S.-H. (2021). Revisiting internal covariate shift for batch normalization. IEEE Transactions on Neural Networks and Learning Systems, 32(11), 5082– 5092. https://doi.org/10.1109/tnnls.2020.3026784

4. Aydemir, O., & Kayikcioglu, T. (2014). Classification of Electroencephalogram signals based on cursor movement imagery. 2014 22nd Signal Processing and Communications Applications Conference (SIU). https://doi.org/10.1109/siu.2014.6830365

5. Aydin, E. A. (2022). Classification of forearm movements by using movement related cortical potentials. 2022 Innovations in Intelligent Systems and Applications Conference (ASYU). https://doi.org/10.1109/asyu56188.2022.9925301

6. Barakat, A., & Bianchi, P. (2021). Convergence and dynamical behavior of the Adam Algorithm for nonconvex stochastic optimization. SIAM Journal on Optimization, 31(1), 244–274. https://doi.org/10.1137/19m1263443

7. Baumgartner, C. F., Oktay, O., & Rueckert, D. (2017). Fully convolutional networks in medical imaging: Applications to image enhancement and recognition. Deep Learning and Convolutional Neural Networks for Medical Image Computing, 159–179. https://doi.org/10.1007/978-3-319-42999-1_10

8. Bekiryazici, S., Demir, A., & Yilmaz, G. (2020). Feature selection and analysis EEG signals with sequential forward selection algorithm and different classifiers. 2020 28th Signal Processing and Communications Applications Conference (SIU). https://doi.org/10.1109/siu49456.2020.9302482

9. Bengio, Y., Lamblin, P., Popovici, D., & Larochelle, H. (2007). Greedy layer-wise training of Deep Networks. Advances in Neural Information Processing Systems 19, 153–160. https://doi.org/10.7551/mitpress/7503.003.0024

10. Friedman, J. H. (2001). Greedy function approximation: A gradient boosting machine. The Annals of Statistics, 29(5). https://doi.org/10.1214/aos/1013203451

11. Hernandez-Galvan, A., Ramirez-Alonso, G., Camarillo-Cisneros, J., Samano-Lira, G., & Ramirez-Quintana, J. (2022). Imagined speech recognition in a subject independent approach using a prototypical network. IFMBE Proceedings, 37–45. https://doi.org/10.1007/978-3-031-18256-3_4

12. Jiang, G., & Wang, W. (2017). Error estimation based on variance analysis of K -fold cross-validation. Pattern Recognition, 69, 94–106. https://doi.org/10.1016/j.patcog.2017.03.025

13. Karakaya, S., Kucukyildiz, G., & Ocak, H. (2017). Classification of motor imaginary in EEG using random. Global Journal of Computer Sciences: Theory and Research, 7(3), 129–135. https://doi.org/10.18844/gjcs.v7i3.2792

14. Kareem Abdullah, A., & Chao Zhu, Z. (2014). Blind source separation based of Brain Computer Interface System: A Review. Research Journal of Applied Sciences, Engineering and Technology, 7(3), 484–494. https://doi.org/10.19026/rjaset.7.280

15. Khademi, Z., Ebrahimi, F., & Kordy, H. M. (2022). A transfer learning-based CNN and LSTM hybrid deep learning model to classify motor imagery EEG signals. Computers in Biology and Medicine, 143, 105288. https://doi.org/10.1016/j.compbiomed.2022.105288

16. Krizhevsky, A., Sutskever, I., & Hinton, G. E. (2017). ImageNet classification with deep convolutional Neural Networks. Communications of the ACM, 60(6), 84–90. https://doi.org/10.1145/3065386

17. Lecun, Y., Bottou, L., Bengio, Y., & Haffner, P. (1998). Gradient-based learning applied to document recognition. Proceedings of the IEEE, 86(11), 2278–2324. https://doi.org/10.1109/5.726791

18. Li, J., Zhang, H., Zhao, J., Guo, X., Rihan, W., & Deng, G. (2022). Embedded feature selection and machine learning methods for flash flood susceptibility-mapping in the mainstream Songhua River Basin, China. Remote Sensing, 14(21), 5523. https://doi.org/10.3390/rs14215523

19. Lopez-Larraz, E., Antelis, J. M., Montesano, L., Gil-Agudo, A., & Minguez, J. (2012). Continuous decoding of motor attempt and motor imagery from EEG activity in spinal cord injury patients. 2012 Annual International Conference of the IEEE Engineering in Medicine and Biology Society. https://doi.org/10.1109/embc.2012.6346299

20. Machado, M. R., Karray, S., & de Sousa, I. T. (2019). LIGHTGBM: An effective decision tree gradient boosting method to predict customer loyalty in the finance industry. 2019 14th International Conference on Computer Science & Education (ICCSE). https://doi.org/10.1109/iccse.2019.8845529

21. Martin, J. C., Liley, D. T., Harvey, A. S., Kuhlmann, L., Sleigh, J. W., & Davidson, A. J. (2014). Alterations in the functional connectivity of frontal lobe networks preceding emergence delirium in children. Anesthesiology, 121(4), 740–752. https://doi.org/10.1097/aln.0000000000000376

22. Mudgal, S. K., Sharma, S. K., Chaturvedi, J., & Sharma, A. (2020). Brain Computer Interface Advancement in Neurosciences: Applications and Issues. Interdisciplinary Neurosurgery, 20, 100694. https://doi.org/10.1016/j.inat.2020.100694

23. Mwata-Velu, T., Ruiz-Pinales, J., Rostro-Gonzalez, H., Ibarra-Manzano, M. A., Cruz-Duarte, J. M., & Avina-Cervantes, J. G. (2021). Motor imagery classification based on a recurrent-convolutional architecture to control a hexapod robot. Mathematics, 9(6), 606. https://doi.org/10.3390/math9060606

24. Ofner, P., Schwarz, A., Pereira, J., Wyss, D., Wildburger, R., & Müller-Putz, G. R. (2019). Attempted arm and hand movements can be decoded from low-frequency EEG from persons with Spinal Cord Injury. Scientific Reports, 9(1). https://doi.org/10.1038/s41598-019-43594-9

25. Peng, J. (2018). Understanding of the convolutional neural networks with relative learning algorithms. 3rd International Conference on Electromechanical Control Technology and Transportation. https://doi.org/10.5220/0006976406570661

26. Poernomo, A., & Kang, D.-K. (2018). Biased dropout and Crossmap dropout: Learning towards effective dropout regularization in Convolutional Neural Network. Neural Networks, 104, 60–67. https://doi.org/10.1016/j.neunet.2018.03.016

27. Primartha, R., Adhi Tama, B., Arliansyah, A., & Januar Miraswan, K. (2019). Decision tree combined with PSO-based feature selection for sentiment analysis. Journal of Physics: Conference Series, 1196, 012018. https://doi.org/10.1088/1742-6596/1196/1/012018

28. Raihan, Md. J., Khan, Md. A.-M., Kee, S.-H., & Nahid, A.-A. (2023). Detection of the chronic kidney disease using XGBoost classifier and explaining the influence of the attributes on the model using shap. Scientific Reports, 13(1). https://doi.org/10.1038/s41598-023-33525-0

29. Robinson, N., Mane, R., Chouhan, T., & Guan, C. (2021). Emerging trends in BCI-robotics for motor control and Rehabilitation. Current Opinion in Biomedical Engineering, 20, 100354. https://doi.org/10.1016/j.cobme.2021.100354

30. Stone, M. (1974). Cross-validatory choice and assessment of statistical predictions. Journal of the Royal Statistical Society: Series B (Methodological), 36(2), 111–133. https://doi.org/10.1111/j.2517-6161.1974.tb00994.x

31. Tariq, M., Trivailo, P. M., & Simic, M. (2018). EEG-based BCI control schemes for lower-limb assistive-robots. Frontiers in Human Neuroscience, 12. https://doi.org/10.3389/fnhum.2018.00312

32. Tayeb, Z., Fedjaev, J., Ghaboosi, N., Richter, C., Everding, L., Qu, X., Wu, Y., Cheng, G., & Conradt, J. (2019). Validating deep neural networks for online decoding of motor imagery movements from EEG signals. Sensors, 19(1), 210. https://doi.org/10.3390/s19010210

33. Torres, E. P., Torres, E. A., Hernández-Álvarez, M., & Yoo, S. G. (2020). EEG-based BCI EMOTION RECOGNITION: A survey. Sensors, 20(18), 5083. https://doi.org/10.3390/s20185083

34. Uyulan, C. Development of LSTM&CNN based hybrid deep learning model to classify motor imagery tasks. (2021). Communications in Mathematical Biology and Neuroscience. https://doi.org/10.28919/cmbn/5265

35. Varma, S., & Simon, R. (2006). Bias in error estimation when using cross-validation for model selection. BMC Bioinformatics, 7(1). https://doi.org/10.1186/1471-2105-7-91

36. Wang, C., Deng, C., & Wang, S. (2020). Imbalance-xgboost: Leveraging weighted and focal losses for binary label-imbalanced classification with XGBoost. Pattern Recognition Letters, 136, 190–197. https://doi.org/10.1016/j.patrec.2020.05.035

37. Wei, L., Yue, H., Jiang, X., Xi, C., & Xiaojun, W. (2009). The assessment of EEG in patients with spinal cord injury to movements. 2009 Third International Symposium on Intelligent Information Technology Application Workshops. https://doi.org/10.1109/iitaw.2009.15

38. Xu, F., Li, J., Dong, G., Li, J., Chen, X., Zhu, J., Hu, J., Zhang, Y., Yue, S., Wen, D., & Leng, J. (2022). EEG decoding method based on multi-feature information fusion for Spinal Cord Injury. Neural Networks, 156, 135–151. https://doi.org/10.1016/j.neunet.2022.09.016

39. Zhang, G., Davoodnia, V., Sepas-Moghaddam, A., Zhang, Y., & Etemad, A. (2020). Classification of hand movements from EEG using a deep attention-based LSTM network. IEEE Sensors Journal, 20(6), 3113–3122. https://doi.org/10.1109/jsen.2019.2956998

40. Zhang, R., Zong, Q., Dou, L., & Zhao, X. (2019). A novel hybrid deep learning scheme for four-class motor imagery classification. Journal of Neural Engineering, 16(6), 066004. https://doi.org/10.1088/1741-2552/ab3471

41. Zhang, Y., Tan, X., & GU, Y. (2022). A CNN-LSTM network for classification of attention deficit hyperactivity disorder from EEG Data. 2022 41st Chinese Control Conference (CCC). https://doi.org/10.23919/ccc55666.2022.9902112

